# Applying a Motivational Instructional Design Model to Stroke Rehabilitation: A Feasibility Study in Occupational Therapy and Swallowing Therapy Settings

**DOI:** 10.1101/2024.01.10.24301094

**Authors:** Kazuaki Oyake, Shota Watanabe, Ayano Takeuchi, Taiki Yoshida, Takashi Shigematsu, Yuuki Natsume, Shigeki Tsuzuku, Kunitsugu Kondo, Ichiro Fujishima, Yohei Otaka, Satoshi Tanaka

## Abstract

**OBJECTIVE:** This feasibility study aimed to investigate the feasibility of applying a motivational instructional design model to stroke rehabilitation and its potential physical and mental health effects in occupational and swallowing therapy settings.

**DESIGN:** An open-label, single-arm, feasibility study.

**SETTING:** Convalescent rehabilitation hospitals.

**PARTICIPANTS:** Twenty-five patients with stroke (19 males; mean age 62.4 ± 11.9 years) were recruited from two convalescent rehabilitation hospitals.

**INTERVENTIONS:** The intervention was to motivate participants during rehabilitation sessions based on the motivational instructional design model and was delivered to 12 and 13 participants in two hospitals during occupational and swallowing therapy sessions, respectively. The intervention was given for 40–60 min daily, 5 days weekly for 4 weeks (25 sessions).

**MAIN OUTCOME MEASURES:** The primary outcome was feasibility, including the drop-out rate, an adverse event, and the participants’ acceptability of the intervention. Additionally, physical (activities of daily living, motor function of the paretic upper extremity, and swallowing ability) and mental health (depressive symptoms and apathy) outcomes were evaluated before and after the intervention.

**RESULTS:** No participants dropped out of the intervention or experienced an adverse event. Twenty-one participants (84%) were satisfied with the intervention, and 19 (76%) hoped to continue receiving it. After the intervention, statistically significant improvements with a large effect size were found in physical outcomes (Cohen’s r = 0.68–0.85) but not in mental health outcomes (Cohen’s r = 0.31–0.34).

**CONCLUSIONS:** The application of the motivational instructional design model to occupational and swallowing therapies after stroke was feasible with the potential to improve physical outcomes.

## INTRODUCTION

Stroke is a primary cause of disability and requires continuous care.^1^ Rehabilitation is recommended to promote functional recovery, enhance independence in daily activities, and improve quality of life after a stroke.^2^ For example, occupational therapy, including task-specific training and functional task practice, has been demonstrated to improve the motor function of the paretic upper extremity and activities of daily living.^2, 3^ In addition, swallowing exercises, including tongue exercise and effortful swallowing, improve dysphagia after stroke.^4, 5^ The independent efforts of the patient are essential to sustain these practices and exercises. Therefore, motivation, a “mental function that produces the incentive to act; the conscious or unconscious driving force for action,”^6^ may be associated with improved functional recovery after stroke.^7-9^

Motivation for rehabilitation programs is a dynamic condition rather than a static quality.^10^ Social factors and the personality or clinical characteristics of the patient are possible determinants of motivation for rehabilitation.^10-13^ For example, negative experiences and rehabilitation perceptions, such as boredom, low self-efficacy, and inadequate knowledge about the benefits of exercise, may decrease motivation.^10-12^ Although various motivational interventions, such as motivational interviewing and goal setting, are used in stroke rehabilitation,^14-20^ there is insufficient evidence indicating that these interventions contribute to improvements in rehabilitation outcomes.^21, 22^ Therefore, developing an additional adjuvant strategy to improve patient motivation and outcomes is essential.

The Attention, Relevance, Confidence, and Satisfaction (ARCS) model is a motivational instructional design model that structurally presents policies and procedures on how educators should design learning environments to motivate students to learn based on motivational theories.^23, 24^ Similarly, it helps educators to identify the component of instruction that affect student motivation and to determine strategies for solving motivational challenges in instructional materials and methods. The ARCS model is currently applied to various fields such as engineering, pharmacy, and nursing.^25-30^ Regarding rehabilitation, only one study has involved using the ARCS model for patient education for fall prevention.^25^ The ARCS model can help therapists select the appropriate motivational strategies according to the cause of a patient’s lack of motivation, which may effectively motivate patients to engage in rehabilitation.^17, 20^ However, to our knowledge, no study has involved applying this model to motivate patients during rehabilitation programs, including functional training and task practice. Thus, we aimed to investigate the feasibility of applying the motivational instructional design model to stroke rehabilitation and its potential effects on physical and mental health outcomes in occupational and swallowing therapy settings.

## METHODS

### STUDY DESIGN

This was an open-label, single-arm, feasibility study. The study protocol was approved by the appropriate ethics committees at the two hospitals (approval numbers: 216-2 and 18-62). All participants provided written informed consent before enrollment in the study. The study was conducted according to the Declaration of Helsinki of 1964, as revised in 2013. The study protocol was pre-registered in the University Hospital Medical Information Network (UMIN 000037324 and UMIN000037506).

### PARTICIPANTS

Patients hospitalized in a convalescent rehabilitation ward were consecutively recruited from two hospitals between August 2019 and March 2022. Patients in one of the two hospitals received an intervention described below during occupational therapy sessions (OT group), and those in another hospital underwent the intervention during swallowing therapy sessions (ST group). The inclusion criteria common to participants in both groups comprised the following: age 40–90 years; being within 180 days of first-ever stroke; having received occupational or swallowing therapy for at least 1 week after admission; scheduled to be hospitalized for at least 4 weeks in the future during screening; a Mini-Mental State Examination score of at least 24 points.^31^ An additional inclusion criterion for participants in the OT group was having unilateral upper extremity motor paralysis defined using an upper extremity motor subscale of Fugl-Meyer Assessment score of less than 66 points,^32^ and that for those in the ST group was having dysphagia defined using a Food intake LEVEL Scale (FILS) score of less than 9 points.^33^ Participants were excluded if they had mental impairment hindering their compliance or any comorbid neurological disorders. Demographic and clinical data, such as age and stroke type, were obtained from patient medical records.

### INTERVENTION

The intervention was to explicitly motivate participants during inpatient daily rehabilitation sessions based on the ARCS model. All participants underwent one-on-one conventional inpatient daily rehabilitation: 40–60 min of physical and 40–60 min of occupational therapies. Additionally, participants with dysphagia received 40 min of swallowing therapy, 5 days weekly. In the OT group, motivational strategies based on the ARCS model were implemented in 5 days of occupational therapy weekly. In the ST group, the motivational strategies were also implemented in 5 days of swallowing therapy sessions weekly.

The ARCS-based approach comprised an iterative process with a cycle of assessment of the patient’s motivational challenges, design of motivational strategies, and implementation of motivational strategies (**Figure 1**). Each detail is described below.

**Figure 1.**
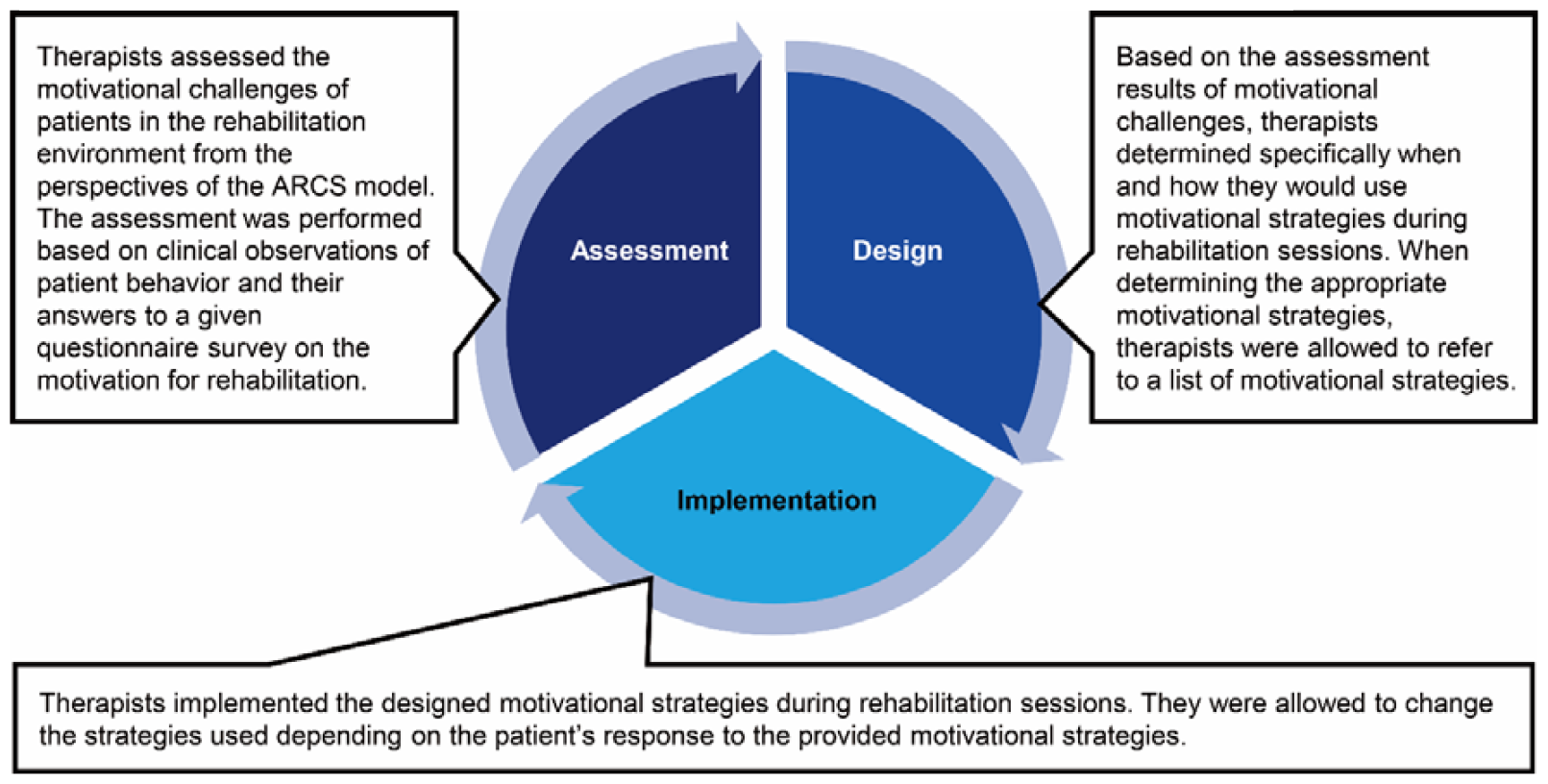
Intervention involving the application of the ARCS model of motivational instructional design to rehabilitation. ARCS: Attention, Relevance, Confidence, and Satisfaction.

#### Assessment of motivational challenges

Before the intervention, therapists assessed the motivational challenges of patients in the rehabilitation environment from the following four perspectives: (1) direction of attention and interest in the provided rehabilitation programs, (2) understanding of the relevance of the provided rehabilitation programs to their goals and needs, (3) confident accomplishment of the provided rehabilitation programs, and (4) satisfaction with the provided rehabilitation programs. The assessment was performed based on clinical observations of patient behavior and their answers to a given questionnaire survey on the motivation for rehabilitation. The survey included four items corresponding to the four dimensions of the ARCS model. Each item was rated on a 5-point Likert scale that ranges from 1 to 5 (1 = strongly disagree, 2 = disagree, 3 = neither agree nor disagree, 4 = agree, 5 = strongly agree). In addition, the sum of the scores of the four items was calculated as the motivation level score. The survey given to participants in the OT group is shown in **Box 1**. At the end of each of the 5 sessions, the motivational challenges were reassessed to modify the design of motivational strategies.

##### Box 1.

Survey presented to participants in the OT group

Below are statements regarding your experiences and perceptions of occupational therapy programs provided by me within the past seven days from now. Please read each one and indicate to what extent you agree or disagree with each statement.

Statement 1: I was interested in the provided occupational therapy programs.

○ strongly disagree ○ disagree ○ neither agree nor disagree ○ agree ○ strongly agree

Statement 2: I understood the relevance of the provided occupational therapy programs to my goals and needs.

○ strongly disagree ○ disagree ○ neither agree nor disagree ○ agree ○ strongly agree

Statement 3: I had the confidence to accomplish the provided occupational therapy programs.

○ strongly disagree ○ disagree ○ neither agree nor disagree ○ agree ○ strongly agree

Statement 4: I was satisfied with the provided occupational therapy programs.

○strongly disagree ○ disagree ○ neither agree nor disagree ○ agree ○ strongly agree

Participants in the ST group were given a survey in which “occupational therapy” was replaced by “swallowing therapy.”

Abbreviations: OT, occupational therapy; ST, swallowing therapy.

#### Design of motivational strategies

Based on the assessment results of motivational challenges, therapists determined specifically when and how they would use motivational strategies during rehabilitation sessions. In this study, motivational strategies were defined as concrete tactics, techniques, or approaches to orient patients to rehabilitation by solving motivational challenges.^18^ When determining the appropriate motivational strategies, therapists were allowed to refer to a list of motivational strategies (**Table 1**). The list was initially developed by the first author based on data obtained from the educational literature related to the ARCS model,^23, 24, 26, 30, 34^ semi-structured interviews with physical therapists,^35^ and our previous study on motivational strategies for stroke rehabilitation.^19^ Finally, the first and last authors who learned about the ARCS model completed the list after reviewing the contents for clarity and relevance.

**Table 1.**
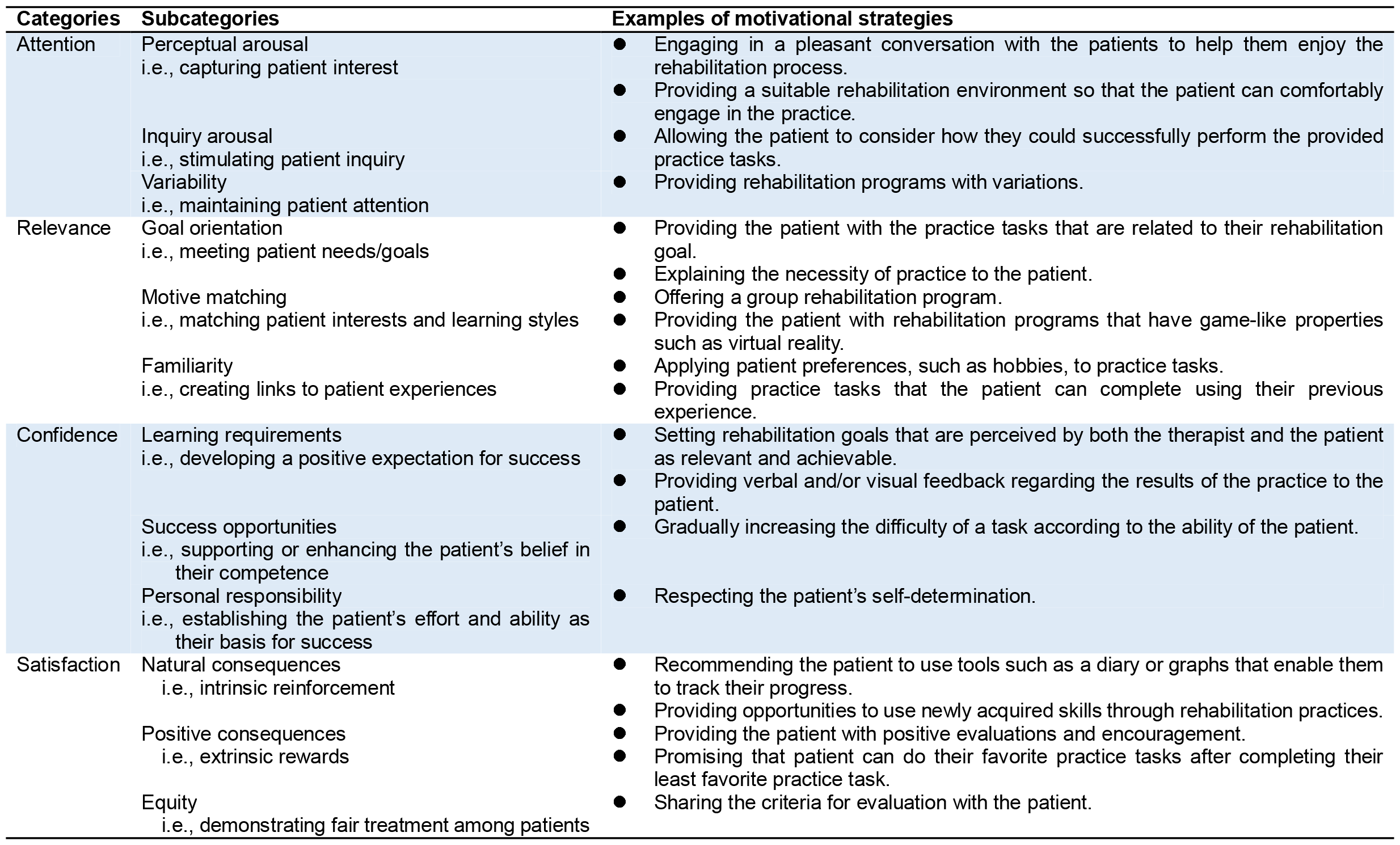
List of motivational strategies based on the Attention, Relevance, Confidence, and Satisfaction model.

Some examples of the design of motivational strategies according to the motivational challenges are given below. If a patient rated with a score of 1 (strongly disagree) for the item “*I understand the relevance of the provided rehabilitation programs to my goals and needs*” of the survey, the therapist would explain the expected training benefits to the patient before providing it. In contrast, when a patient seemed to be bored during the practice despite a score of 5 (strongly agree) for all items of the questionnaire, the therapist would change rehabilitation programs daily to prevent the patient from getting bored. Furthermore, even if a patient seemed to be actively engaging in rehabilitation programs and rated with a score of 5 for all items of the questionnaire, the therapist was asked to use the potential strategies to maintain the patient’s motivation.

#### Implementation of motivational strategies

Therapists implemented the designed motivational strategies during rehabilitation sessions. In the OT group, the occupational therapy programs included task-specific practice to improve the motor function of the paretic upper extremity and the activities of daily living training tailored to individual needs. In the ST group, the swallowing therapy programs included indirect and direct training, such as tongue exercise and feeding/swallowing foods and/or liquids. Therapists were allowed to change the strategies used depending on the patient’s response to the provided motivational strategies.

#### Therapist training in delivering the intervention

All therapists received two 60-min training sessions in delivering the intervention by the first and last authors before starting participant enrollment. On the first day of training, they learned the concept of the ARCS model. On the second day of training, they practiced procedures for determining motivational strategies using the ARCS model. To ensure whether the intervention was appropriately implemented during the intervention period, they were asked to report on the design and implementation of motivational strategies in the intervention over the past week to the researchers via email weekly. Based on the reports from therapists, the researchers provided them with corrective feedback as needed.

### OUTCOME MEASURES

The primary outcome was feasibility, including the drop-out rate, an adverse event, and the participants’ acceptability of the intervention.^36, 37^ In addition, physical and mental health outcomes were evaluated before and after the intervention. The assessments of these outcomes were performed by the same therapists who provided the participants with the intervention.

#### Primary outcome

Therapists recorded the occurrence of drop-outs and an adverse event during the intervention period. To assess the acceptability of the intervention, the participants were asked to rate the two items on satisfaction with the intervention and intention to continue receiving it using a 5-point Likert scale ranging from “strongly disagree” to “strongly agree” after the intervention (**Box 2**). For quantitative assessment of acceptability, our threshold was that at least 75% of the participants answered “agree” or “strongly agree” to the two items.^38^

#### Physical and mental health outcomes

P Physical outcomes included activities of daily living, motor function of the paretic upper extremity, and swallowing ability. The degrees of independence in activities of daily living were evaluated with the motor subscale of the Functional Independence Measure (FIM).^39^ In the OT group, the motor function of the paretic upper extremity was assessed using the Motor Activity Log amount of use and quality of movement scales.^40^ In the ST group, swallowing ability was evaluated using the Mann Assessment of Swallowing Ability (MASA) and the FILS.^33, 41^ Mental health outcomes included depressive symptoms and apathy, which were assessed using the Self-rating Depression Scale (SDS)^42^ and the Apathy Scale^43^, respectively. The raw sum score of the SDS ranges from 20 to 80 points, and a score of ≥50 points indicated the presence of depressive symptoms.^44^ The total score of the Apathy Scale ranges from 0 to 42 points, and participants with a score of ≥14 points were classified as having an apathy.^43^

##### Box 2.

Survey to assess the participants’ acceptability of the intervention

Below are statements regarding your acceptability of the intervention provided in this study. Please read each one and indicate to what extent you agree or disagree with each statement.

Statement 1: I am satisfied with the intervention.

○ strongly disagree ○ disagree ○ neither agree nor disagree ○ agree ○ strongly agree

Statement 2: I hope to continue receiving the intervention.

○ strongly disagree ○ disagree ○ neither agree nor disagree ○ agree ○ strongly agree

### STATISTICAL ANALYSES

There is little consensus on the appropriate sample size for a feasibility study.^45^

According to a previous study,^46^ a sample of 24 participants (12 per group) was required to provide useful information about the feasibility aspects of the study.

We used descriptive statistics to characterize the study sample and summarize the feasibility outcomes. Demographic and clinical characteristics were compared between the OT and ST groups using the unpaired t-test for continuous variables and Fisher’s exact test for dichotomous variables. Additionally, the Friedman test was used to examine changes in the motivation level score during the intervention period. To compare physical and mental health outcomes before and after the intervention, we used the Wilcoxon signed-rank test and calculated its Cohen’s *r* as a measure of the effect size, where 0.1 is considered a small effect size; 0.3, a medium effect size; and 0.5, a large effect size.^47^ Statistical analyses were performed using the Statistical Package for the Social Sciences software, version 27.0.^a^ P<0.05 was considered statistically significant.

## RESULTS

### PARTICIPANTS

Twenty-five patients with stroke participated in this study, 12 of whom were in the OT group, and 13 in the ST group. The demographic characteristics of the participants are presented in Table 2. No significant differences were observed in the participants’ characteristics between the groups.

**Table 2.** Participants’ characteristics.

| Variable | Overall<br>(n = 25) | OT group<br>(n = 12) | ST group<br>(n = 13) | p-value |
| --- | --- | --- | --- | --- |
| Age (years) | 62.4 ± 11.9 | 61.2 ± 13.3 | 63.6 ± 11.1 | 0.620 |
| Sex, male/female | 19/6 | 9/3 | 10/3 | 0.999 |
| Type of stroke, ischemic/hemorrhagic | 14/11 | 7/5 | 7/6 | 0.999 |
| Side of motor paresis, right/left | 12/13 | 6/6 | 6/7 | 0.999 |
| Time since stroke (days) | 61.9 ± 36.8 | 65.2 ± 29.7 | 58.8 ± 43.4 | 0.677 |

Values are presented as mean ± standard deviation and number.
P values indicate significant differences between the OT and ST groups.
Abbreviations: OT, occupational therapy; ST, swallowing therapy.

### FEASIBILITY OF THE INTERVENTION

No participants dropped out or experienced an adverse event during the intervention period. The results of the survey regarding the acceptability of the intervention are shown in **Figure 2**. Twenty-one (84%) participants responded with “agree” or “strongly agree” to the statement regarding their satisfaction with the intervention (**Figure 2A**). Additionally, 19 (76%) participants answered “agree” or “strongly agree” to the item regarding their intention to continue receiving the intervention (**Figure 2B**). These results indicate a good acceptability of the intervention. The survey on the motivation for rehabilitation was administered to all participants every week. The median motivation level score significantly increased during the intervention period (χ^2^ = 22.23, p < 0.001: **Figure 3**).

**Figure 2.**
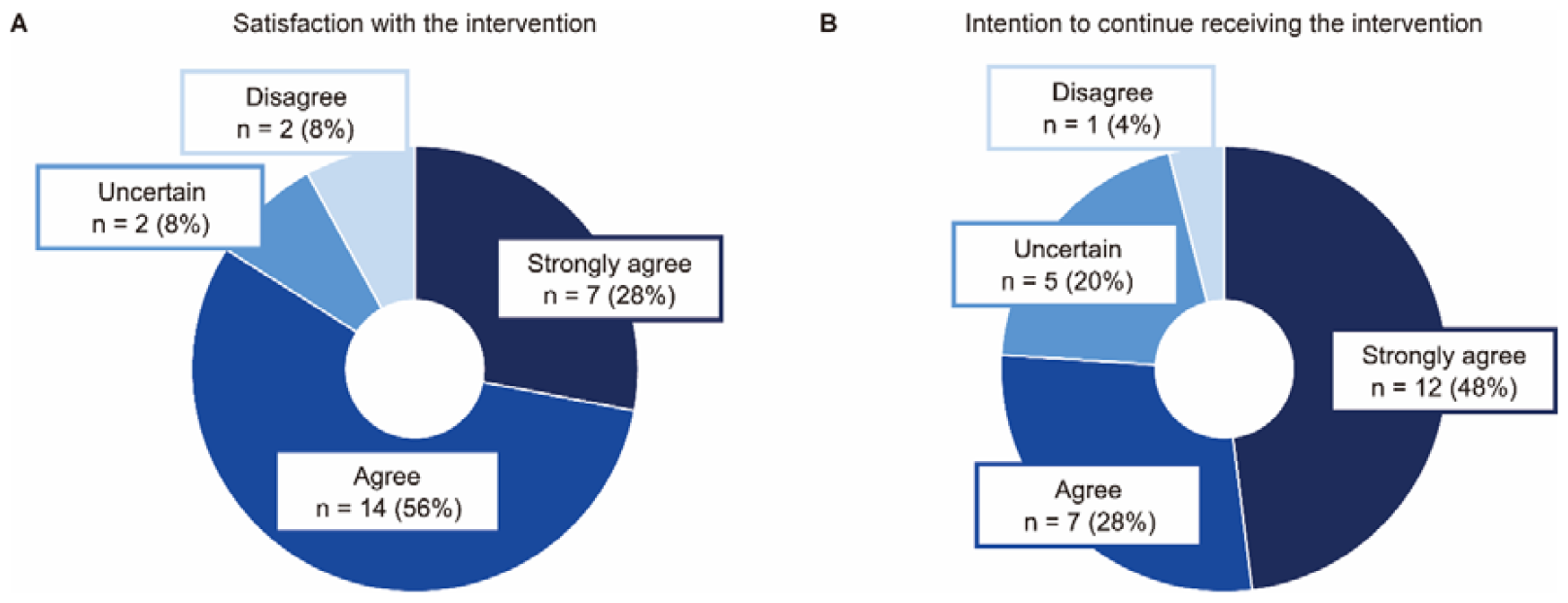
Participants’ responses to a survey on the acceptability of the intervention. (A) The result of the survey regarding the satisfaction with the intervention. (B) The result of the survey regarding the intention to continue receiving the intervention.

**Figure 3.**
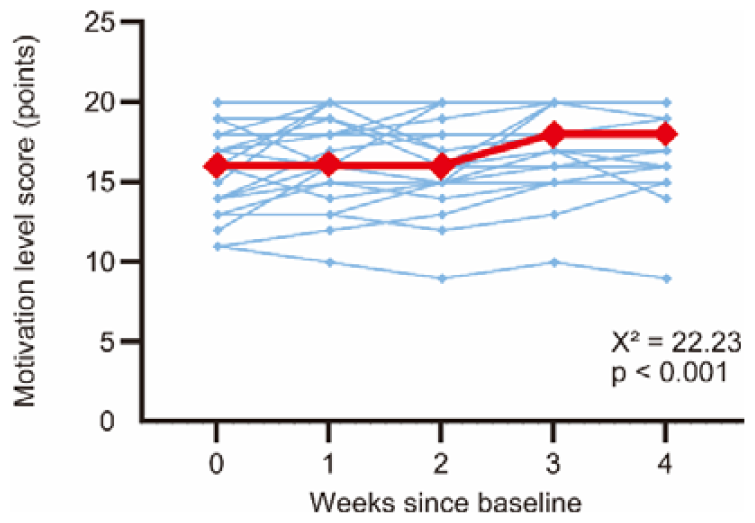
Changes in the motivation level score during the intervention. The red line represents the median data. Blue lines represent the individual participant data.

### POTENTIAL PHYSICAL AND MENTAL HEALTH EFFECTS OF THE INTERVENTION

Changes in physical outcomes after the intervention compared to before are shown in **Figure 4**. Statistically significant improvements with a large effect size were observed in the scores of the FIM motor items (p < 0.001, Cohen’s r = 0.80: **Figure 4A**), the Motor Activity Log amount of use (p = 0.006, Cohen’s r = 0.80: **Figure 4B**), and quality of movement scales (p = 0.018, Cohen’s r = 0.68: **Figure 4C**), the MASA (p = 0.003, Cohen’s r = 0.85: **Figure 4D**), and the FILS (p = 0.010, Cohen’s r = 0.72: **Figure 4E**). The differences in the SDS (p = 0.124, Cohen’s r = 0.31: **Figure 5A**) and Apathy Scale (p = 0.091, Cohen’s r = 0.34: **Figure 5B**) scores before and after the intervention were not statistically significant despite a medium effect size. However, before the intervention, five and 12 participants had scores of ≥50 points and ≥14 points on the SDS and Apathy Scale, respectively. Thus, we included only these participants in the subgroup analyses. In five participants with depressive symptoms before the intervention, the median [interquartile range] SDS score decreased from before (54 [52–55] points) to after the intervention (46 [45–52] points), with a large effect size (p = 0.125, Cohen’s r = 0.78). Additionally, in 12 participants with apathy before the intervention, the median [interquartile range] Apathy Scale score statistically significantly decreased from before (18.5 [16.75–27.00] points) to after the intervention (16.00 [13.00–23.75] points), with a large effect size (p = 0.007, Cohen’s r = 0.75).

**Figure 4.**
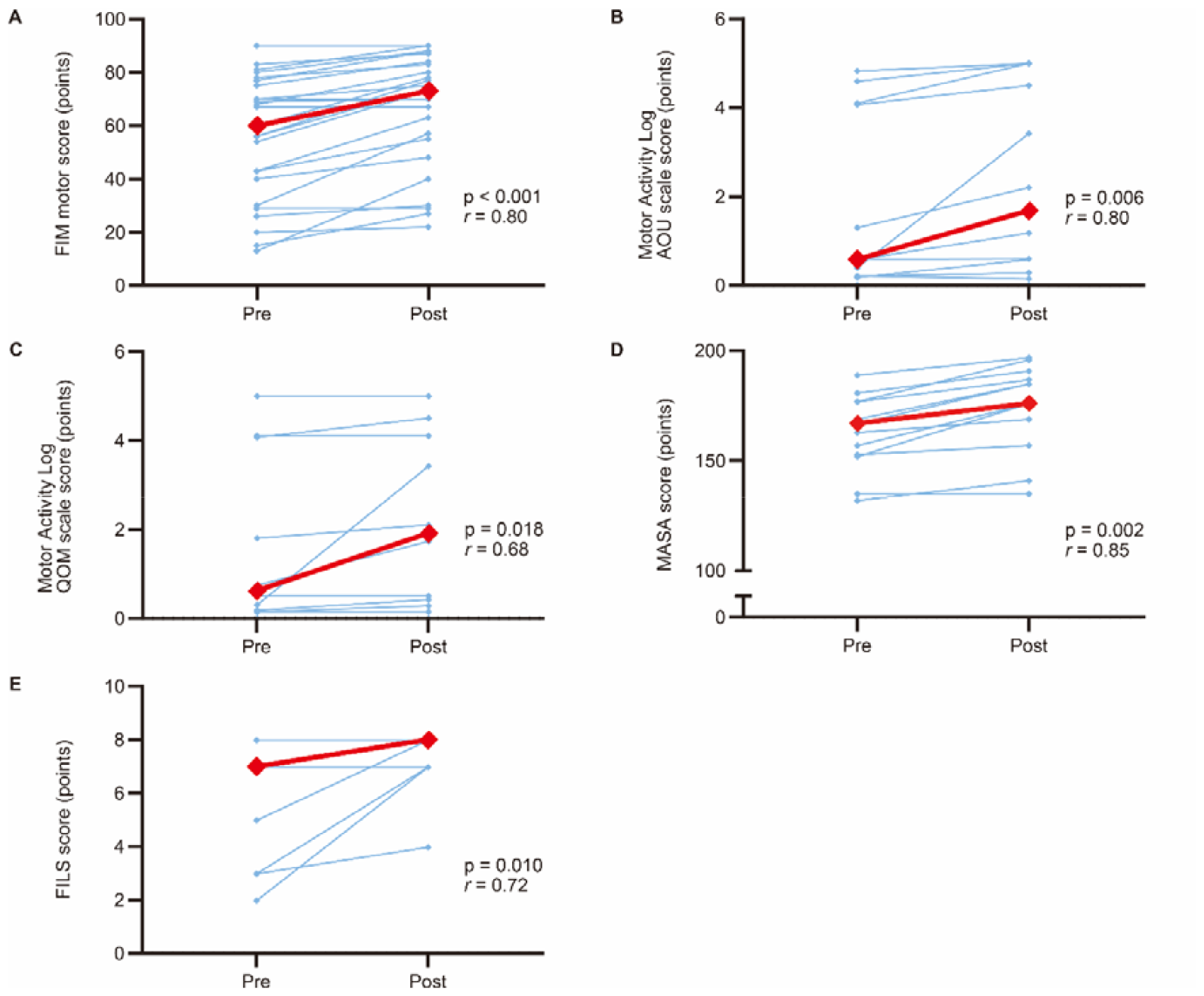
Changes in physical outcomes before and after the intervention. (A) The FIM motor score. (B) The Motor Activity Log AOU scale score. (C) The Motor Activity Log QOM scale score. (D) The MASA score. (E) The FILS score. Red lines represent the median data. Blue lines represent the individual participant data. r: Cohen’s r for the Wilcoxon signed-rank test; FIM: Functional Independence Measure; AOU: amount of use; QOM: quality of movement; MASA: Mann Assessment of Swallowing Ability; FILS: Food Intake LEVEL Scale.

**Figure 5.**
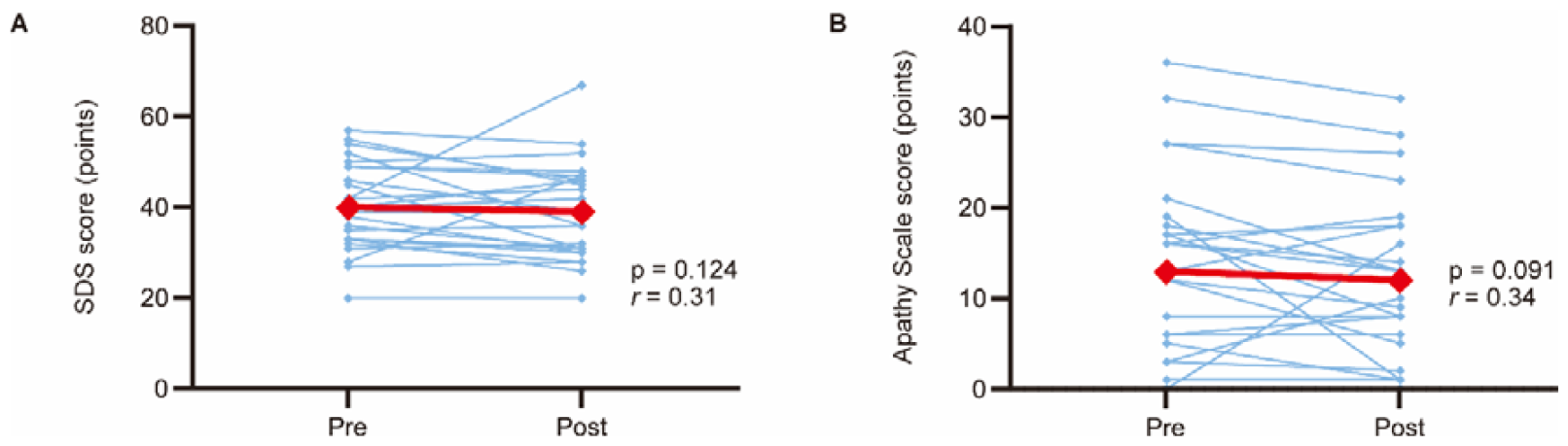
Changes in mental health outcomes before and after the intervention. (A) The SDS score. (B) The Apathy Scale score. Red lines represent the median data. Blue lines represent the individual participant data. r: Cohen’s r for the Wilcoxon signed-rank test; SDS: Self-rating Depression Scale.

## DISCUSSION

The ARCS model is used to design the motivational aspects of learning environments for enhancing and sustaining students’ motivation to learn.^23, 24^ To our knowledge, this study is the first to involve the model in stroke rehabilitation. We demonstrated that applying the ARCS model to occupational and swallowing therapies was feasible. In addition, following the intervention, there were significant improvements with a large effect size in the activities of daily living, the motor function of the paretic upper extremity, and the swallowing ability. We also found no statistically significant improvements in the depressive symptoms and apathy, although these effect sizes were medium. However, only five and 12 participants showed depressive symptoms and apathy, respectively, before the intervention. The subgroup analyses limited to these participants revealed that the SDS and the Apathy Scale scores decreased after the intervention compared to those before with large effect sizes. Therefore, the failure to detect statistically significant changes in mental health outcomes before and after the intervention could be attributed to ceiling effects. The findings of this study may provide therapists with helpful insight into effectively motivating patients to engage in stroke rehabilitation and will offer researchers valuable information for designing future randomized controlled trials.^48^

In this study, participants were recruited from two hospitals. Additionally, the feasibility of the intervention was assessed in two different rehabilitation settings. These procedures allow for improved generalizability of our results. Moreover, we monitored weekly whether the intervention was delivered as intended, which may enhance the reliability and validity of the intervention.^49^

A representative motivational intervention that has been used in rehabilitation is motivational interviewing, which is a collaborative communication style that helps patients resolve their ambivalence and strengthen intrinsic motivation for behavioral changes.^14, 15, 21, 50^ Although motivational interviewing can improve the mood in patients with stroke, there has been limited evidence on it improving functional outcomes after stroke.^21^ The ARCS model would present two advantages over motivational interviewing in rehabilitation. First, the motivational intervention based on the ARCS model is specifically designed to address motivational concerns in the rehabilitation environment through modifications in therapist behaviors (e.g., explaining the necessity of practice) and the content of rehabilitation programs (e.g., providing rehabilitation programs with variations). This allows therapists to effectively manipulate potential factors that influence patient motivation. Second, the ARCS model is highly accessible and can be readily adopted by therapists, including those with limited clinical experience. The therapists in this study received only 60-min training sessions for 2 days. In contrast, successful implementation of motivational interviewing prompts training in verbal and nonverbal communication skills.^50^ In a randomized controlled trial study, therapists who used motivational interviewing underwent an extensive training program for 4 days, comprising up to 10 practice sessions until they were deemed competent and confident in the technique.^51^ Hence, the ARCS model can be effectively used with relatively minimal training requirements.

### STUDY LIMITATIONS

This study has some limitations. First, we could not determine whether the improvements in physical outcomes observed in this study were attributed to using the ARCS model because of no control group; however, the positive results observed in this study support the need for a randomized controlled trial to investigate the effectiveness of applying the ARCS model in rehabilitation. Second, we could not assess the feasibility of the intervention in physical therapy settings because of the coronavirus disease 2019 restrictions. Although the motivational instruction based on the ARCS model in physical education for college students has been reported to effectively promote learning cognition and health-related physical fitness,^28^ our findings should be carefully generalized to physical therapy, including physical fitness exercise and gait training. Finally, all participants were recruited from convalescent rehabilitation wards. Patients who participated in this study may have more adherence to their rehabilitation programs than acute stroke survivors and community-dwelling people with chronic stroke.^52^ Further feasibility studies in acute and chronic rehabilitation settings would improve the external validity of our findings.

## CONCLUSIONS

This study demonstrated that applying the motivational instructional design model to occupational and swallowing therapies after stroke was feasible with the potential to improve the activities of daily living, motor function of the paretic upper extremity, and swallowing ability. These findings may provide valuable information to design a future randomized controlled trial.

## Data Availability

The datasets used and/or analyzed during this study cannot be made publicly available due to the need for participant confidentiality. However, they are available from the corresponding author on reasonable request.

## SUPPLIERS

a. Statistical Package for the Social Sciences; IBM Corp.

## ACKNOWLEDGMENT

The authors thank Ayano Hosobuchi, Takahiro Nishiyama, Haruna Kawahara, Ayumi Nakamura, Kosaku Katsumata, Yuta Asada, Shogo Nozaki, and Miho Nanbu at the Tokyo Bay Rehabilitation Hospital, Sakura Terasaka, Kaede Yoshino, Hiroki Ikeda, Miyu Nakamura, and Hiroka Takatsuji at the Hamamatsu City Rehabilitation Hospital, and Kenjiro Kunieda at the Gifu University for their help and support.

This work was supported by a grant from JSPS KAKENHI Grant Number JP20K21752 to S.T. The funding source had no involvement in the study design; collection, analysis and interpretation of data; writing of the report; and the decision to submit the article for publication.

## DEVICE STATUS

The manuscript submitted does not contain information about medical devices.

